# Predicting local control of brain metastases after stereotactic radiotherapy with clinical, radiomics and deep learning features

**DOI:** 10.1101/2024.05.13.24307241

**Authors:** Hemalatha Kanakarajan, Wouter De Baene, Patrick Hanssens, Margriet Sitskoorn

## Abstract

**Background and purpose:** Timely identification of local failure after stereotactic radiotherapy for brain metastases allows for treatment modifications, potentially improving outcomes. Previous studies showed that adding radiomics or Deep Learning (DL) features to clinical features increased Local Control (LC) prediction accuracy. However, no study has integrated radiomics, DL, and clinical features into machine learning algorithms to predict LC. We examined whether a model using all these features achieves better accuracy than models using only a subset.

**Materials and methods:** We collected pre-treatment brain MRIs and clinical data for 129 patients at the Gamma Knife Center of the Elisabeth-TweeSteden Hospital. Radiomics features (extracted using the Python radiomics feature extractor) and DL features (extracted using a 3D ResNet model) were combined with clinical features. Performance of a Random Forest classifier was compared across four models trained with: clinical features only; clinical and radiomics features; clinical and DL features; and clinical, radiomics, and DL features.

**Results:** The prediction model utilizing only clinical variables provided an Area Under the receiver operating characteristic Curve (AUC) of 0.82 and an accuracy of 75.6%. Adding radiomics features increased the AUC to 0.88 and accuracy to 83.3%, while adding DL features resulted in an AUC of 0.86 and accuracy of 78.3%. The best performance came from combining clinical, radiomics, and DL features, achieving an AUC of 0.89 and accuracy of 87%.

**Conclusion:** Integrating radiomics and DL features with clinical characteristics improves LC prediction after stereotactic radiotherapy for brain metastases. These findings demonstrate the potential for early outcome prediction, enabling timely treatment modifications to improve patient management.

**Clinical and Translational Impact Statement:** Our study holds great clinical value, as the increased prediction accuracy can lead to tailored and effective interventions, resulting in better outcomes for brain metastases patients treated with stereotactic radiotherapy.

## Introduction

Metastatic brain tumors represent the most prevalent form of intracranial malignancies [1]. Brain metastases manifest in approximately 20 to 40% of individuals diagnosed with cancer [2, 3]. While any tumor has the potential to spread to the brain, the predominant types include lung cancer, breast cancer, melanoma, and gastrointestinal cancers [1]. The prevalence of brain metastases is increasing [4]. The adoption of sophisticated imaging methods for diagnosis, alongside the implementation of innovative chemotherapeutic approaches for systemic cancer treatment, may contribute to the increased likelihood of finding and developing brain metastases [1].

Currently, the prognosis of patients with brain metastases is poor with median overall survival of a few weeks to months in untreated patients [5]. The survival of patients with brain metastases depends upon prompt diagnosis and treatment efficacy. The standard treatment options are surgical resection and radiotherapy [5]. Surgery is recommended for patients with a single large tumor in a reachable location [6]. The three principal modalities of radiotherapy for brain metastases are Whole-Brain Radiation Therapy (WBRT), Single-fraction Stereotactic Radiosurgery (SRS), and hypo-fractionated Stereotactic Radiotherapy (SRT). WBRT was the main treatment in the past for patients with multiple brain metastases [7]. There has been a shift from WBRT to SRT and SRS due to the adverse effects of WBRT, such as fatigue and cognitive decline [8]. Through SRS, multiple nonparallel beams are converged to deliver a single, high radiation dose to a targeted region whereas SRT delivers multiple, smaller doses of radiation over time. In SRS and SRT, the delivered radiation is confined to the lesion and there is a rapid dose fall-off at the edge of the treatment volume. Since the radiation dose is not delivered to the healthy brain tissue, there is a reduced likelihood of posttreatment neurocognitive decline compared to WBRT [9].

The assessment of Local Control (LC) of brain metastases is an important clinical endpoint. A stable disease after treatment is categorized as LC while a progressive disease indicates a Local Failure (LF) [10]. It may require several months before local changes of the treated lesions become evident on follow-up scans. Considering that the median survival of patients with brain metastases following radiotherapy can range between 5 months and 4 years [11, 12], timely identification of LF subsequent to radiotherapy is crucial as it offers the opportunity for timely tailored treatment modifications, ensuring that patients receive the most effective care and maximizing their chances of a favorable prognosis.

Cancer imaging analysis driven by Artificial Intelligence (AI) has the potential to revolutionize medical practice by revealing previously undisclosed characteristics from routinely obtained medical images [13]. These features can serve as valuable inputs for the development of machine learning models aimed at predicting the treatment response or LC of brain metastases [13]. This is particularly important given the advancement in Graphical Processing Unit (GPU) processing capabilities and the availability of large amounts of training data which have led to a rapid expansion in neural networks and deep learning techniques for regression and classification tasks [14]. Deep learning models have demonstrated significant potential in identifying crucial and unique features within medical image data across a range of applications, including cancer treatment [15, 16, 17]. Deep learning uses artificial neural networks to automatically learn features from raw data. In medical imaging, deep learning methods are applied directly to the images themselves, learning hierarchical representations of the data. Deep learning has been particularly successful in tasks like image classification, object detection, and segmentation [18]. The information extracted by the deep learning models from the tumor images can be used to predict treatment outcome [18, 19, 20]. Jalalifar et al [21] introduced a novel deep learning architecture to predict the outcome of LC in brain metastasis treated with stereotactic radiation therapy using treatment-planning magnetic resonance imaging (MRI) alongside standard clinical attributes [21]. Their findings highlighted that the addition of deep learning features to the clinical features significantly enhanced the prediction accuracy.

Radiomics is another research domain for extracting quantitative features from medical images for different clinical applications [22]. Radiomics focuses on extracting quantitative features from medical images, such as texture, shape and intensity characteristics. These features are then used to characterize tumors or other abnormalities in the images. While both radiomics and deep learning are used in medical imaging, radiomics focuses on extracting handcrafted features (such as the manually delineated tumor segmentations) from images while deep learning learns features directly from raw data using neural networks [18, 20]. Numerous studies have underscored the efficiency of radiomic-based machine learning algorithms in predicting treatment outcomes across different medical conditions [23, 24]. The radiomic-based machine learning algorithms have also been efficiently applied for the prediction of LC of brain metastases after radiotherapy [25, 26, 27]. Karami et al [25] proposed a radiomics framework to predict the LC in patients with brain metastasis treated with SRT. Based on the radiomics features, Kawahara et al [28] proposed a neural network model for predicting the local response of metastatic brain tumor to Gamma Knife Radiosurgery (GKRS). Liao et al [26] and Andrei et al [27] demonstrated the value of combining radiomic features and clinical features to enhance the prediction of brain metastases responses after GKRS. Their findings show that the addition of radiomic features to the clinical features improved the accuracy of the prediction models for LC of brain metastases.

The studies that used either radiomics or deep learning features together with the clinical features to predict LC of brain metastases after SRT showed that the addition of either radiomics or deep learning features increased the prediction accuracy of the models. But to the best of our knowledge, there is no study that combined both radiomics and deep learning features together with clinical features to develop machine learning algorithms to predict LC of brain metastases. A model trained with all these combined features might predict LC with a higher accuracy than the other models trained with a subset of these features, offering a more comprehensive understanding of treatment response, potentially leading to more tailored and effective interventions which may result in improved treatment outcomes, prolonged patient survival, and enhanced quality of life. Hence, the objective of this study was to predict the LC of brain metastases after SRT using the combination of deep learning, radiomics and clinical features.

## Methods

### Data collection

We retrospectively collected the clinical data from 199 brain metastases patients from the Gamma Knife Center of the Elisabeth-TweeSteden Hospital (ETZ) at Tilburg, The Netherlands. This study was approved by the ETZ science office and by the Ethics Review Board at Tilburg University. The patients underwent GKRS at the Gamma Knife Center. After excluding the patients with incomplete data, we included 129 patients. For these 129 patients, pre-treatment contrast-enhanced (with triple dose gadolinium) brain MRIs were collected using a 1.5T Philips Ingenia scanner (Philips Healthcare, Best, The Netherlands) with a T1-weighted sequence (TR/TE: 25/1.86 ms, FOV: 210x210x150, flip angle: 30°, transverse slice orientation, voxel size: 0.82 x 0.82 x 1.5mm). These high-resolution whole-brain planning scans were made as part of clinical care at the Gamma Knife Center of the ETZ between 2015 and 2021. For all patients, the segmentations of the baseline Ground Truth (GT) were manually delineated by expert oncologists and neuroradiologists at ETZ. These 129 patients were randomly divided into 103 patients for training and 26 patients for testing. At ETZ, the FU MRI scans were made at 3, 6, 9, 12, 15, and 21 months after treatment. A tumor was defined as progressive if there was a relative increase in tumor volume on any of these follow-up MRIs compared to pretreatment MRI. A stable tumor after treatment is categorized as LC while a progressive tumor indicates a LF. The pre-processing, feature extraction, model training, and evaluation were performed in Python (version 3.11).

### Preprocessing

As a first preprocessing step, all the MRI scans were registered to standard MNI space using Dartel in SPM12 (Wellcome Trust Center for Neuroimaging, London, UK), implemented in Python using the Nipype (Neuroimaging in Python–Pipelines and Interfaces) software package (version 1.8.6) [29]. The voxel size of the normalized image was set to 1*1*1 mm. For all other normalization configurations, the default values offered by SPM12 were used. One other preprocessing step was to combine the GT labels for patients with more than one brain metastasis in one single GT mask. FSL library (Release 6.0) was used for this integration. Pre-processing was applied to improve the reliability of radiomics and deep learning feature extraction [30].

### Clinical features

The list of clinical factors that we collected from the Gamma Knife Center of ETZ were gender, survival status, diagnosis of brain metastases within 30 days after diagnosis of primary tumor, prior brain treatment, prior SRS, prior WBRT, prior surgery, prior systemic treatment, presence of extracranial metastases, presence of lymph node metastases, presence of seizure, number of metastases at diagnosis, Karnofsky Performance Status score (KPS), occurrence of new metastases after GKRS, presence of extracranial metastases, primary tumor type, age at diagnosis of brain metastases, age at diagnosis of primary tumor, presence of local recurrence, tumor volume and treatment dose. For the treatment dose, we took the average value from the dose range. We extracted the tumor volume from the segmentations of the baseline GT and added it to the clinical data. We took the total tumor volume across the metastases for patients with more than one brain metastasis. The clinical data was converted to a python dataframe.

### Radiomics features

The segment-based radiomics features were extracted from the T1 weighted pre-treatment MRI scans using the radiomics feature extractor of the python radiomics package. The seven groups of features extracted from the Region Of Interest (ROI) of the tumor segmentations were shape-based features (14 features), first-order features (18), Gray Level Cooccurrence Matrix (GLCM) (24) features, Gray Level Dependence Matrix (GLDM) (14) features, Gray Level Run Length Matrix (GLRLM) (16) features, Gray Level Size Zone Matrix (GLSZM) (16) features, and Neighbouring Gray Tone Difference Matrix (NGTDM) (5) features. The resulting 107 radiomics features were considered in this study. The list of radiomics features extracted are listed in the appendix. The mathematical definitions of these radiomics features are given in the Pyradiomics feature documentation (https://pyradiomics.readthedocs.io/en/latest/features.html). The radiomics features were then combined with the clinical features to form a combined python dataframe.

### Deep learning features

A 3D ResNet model [31, 32] pre-trained on the ImageNet challenge dataset [33] was used to extract the deep learning features from the manually segmented masks. Prior to input, the images were rescaled to 256 × 256 × 256 using spline interpolation order 3, improving the accuracy of the model [34]. Additionally, the pixels were also sample-wise scaled between -1 and 1. These preprocessing steps contribute to optimizing the performance of the 3D ResNet model in extracting meaningful features from the images [35].

A 3D convolution was applied on the training data. This convolutional layer was designed with 50 filters and a large kernel size of (7, 7, 7), while also employing a stride of (2, 2, 2) for down sampling. The purpose of this stride is to efficiently reduce the spatial dimensions of the input data, capturing broader information across the dataset while managing computational complexity [36].

To fine-tune this convolutional layer, we applied batch normalization and Rectified Linear Unit (ReLU) activation. Batch normalization helps the model to adapt to our dataset, improving its performance, stability and ability to generalize. ReLU is like a simple on/off switch for a neuron in a neural network. ReLU helps the neural networks learn by letting positive signals pass through unchanged while ignoring negative ones and thus enables neural networks to learn complex patterns effectively. ReLU transforms the features from the images to be compatible with the pretrained model, maintain consistency with the original training, and facilitate efficient gradient propagation during the fine-tuning process [37, 38].

Following this, we incorporated three fine-tuned ResNet blocks into the model. After adding the ResNet blocks, we applied global average pooling to reduce the spatial dimensions to 1x1x1. Finally, a dense layer with softmax activation was added for classification. The deep learning features were extracted using this fine-tuned 3D ResNet model and then combined with the clinical and radiomics features to form a combined python dataframe.

The complete process of pre-processing and feature extraction is summarized in Figure 1.

**Figure 1:**
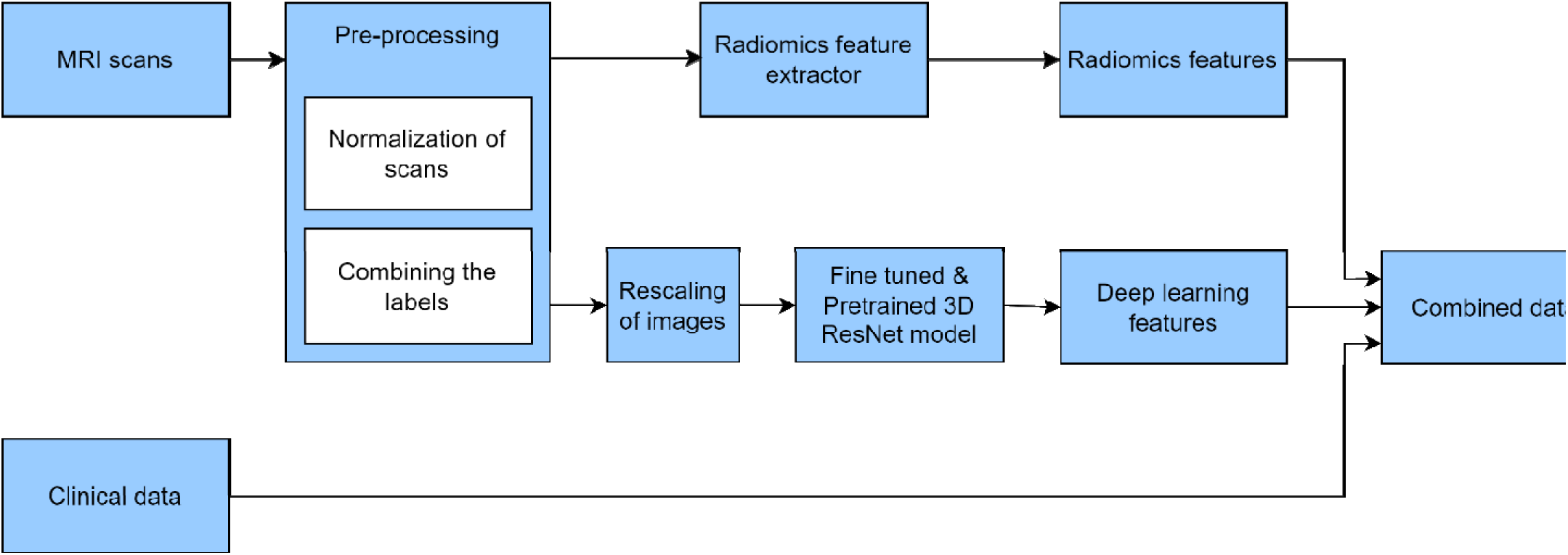
The process of pre-processing, feature extraction, and combining the data.

### Model training

The features with low variance (<0.01) were determined and excluded from the combined dataset to improve prediction accuracy. The list of the excluded features is included in the Appendix. The data was then normalized and was supplied to the Random Forest classifier. Experimental results from Chen et al. [39] demonstrated that the Random Forest machine learning algorithm achieves a better classification performance compared to other classification algorithms. Hence, we choose the Random Forest machine learning algorithm to predict LC from the combined data. The model was trained with the training data set and then tested with the test data set. The process of training and evaluation of the models is shown in Figure 2. The binary outcome used in training and validation was the LC after treatment taken from the list of clinical features. The different models that we created were:

1. Random Forest classifier trained with clinical features only.
2. Random Forest classifier trained with the combination of clinical and deep learning features.
3. Random Forest classifier trained with the combination of clinical and radiomics features.
4. Random Forest classifier trained with the combination of clinical, radiomics and deep learning features.

**Figure 2:**
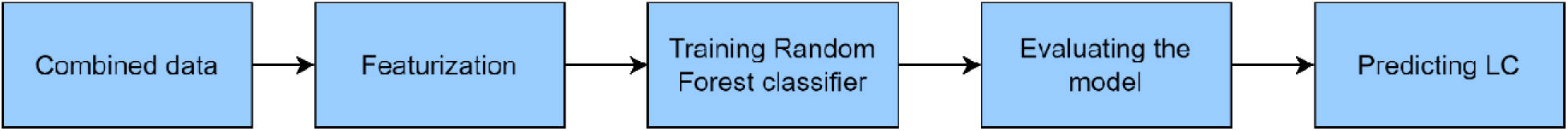
Model training, evaluation and prediction.

### Model evaluation

The performance of the model was evaluated by measuring the following metrics: classification accuracy, precision, F1 score, recall and AUC. The classification accuracy is the ratio of the number of correct predictions to the total number of input samples. The precision is the ability of the classifier not to label as positive a sample that is negative and recall is the ability of the classifier to find all the positive samples. In other words, precision is the ratio of true positive predictions to the total number of positive predictions made by the model, while recall is the ratio of true positive predictions to the total number of actual positives in the dataset. The F1 score in percentage gives the balance between how often the model is correct (precision) and how well it finds all the positive instances (recall). A Receiver Operating Characteristic (ROC) is a graphical plot which is created by plotting the true positive rate vs the false positive rate at various threshold settings. The AUC computes the area under the ROC curve. By doing so, the curve information is summarized in one number. Similar to the F1 score, the AUC reaches its best value at 1.

A K-fold cross validation was applied on the model. The different values that we used for K during cross-validation were 2, 3, and 5. The average accuracy and other metrics across the different folds was calculated. From the trained models, we also extracted the importance of the various factors for predicting the LC.

## Results

### Patient characteristics

Table 1 shows the characteristics of patients included in our study. Among the 129 patients, 42% were male and 58% were female. The patients had an average age of 63 and an average tumor volume of 17,445 mm^3^. Sixty-nine % of the patients had a primary lung cancer and 94% of the patients had less than 10 brain metastases.

**Table 1:**
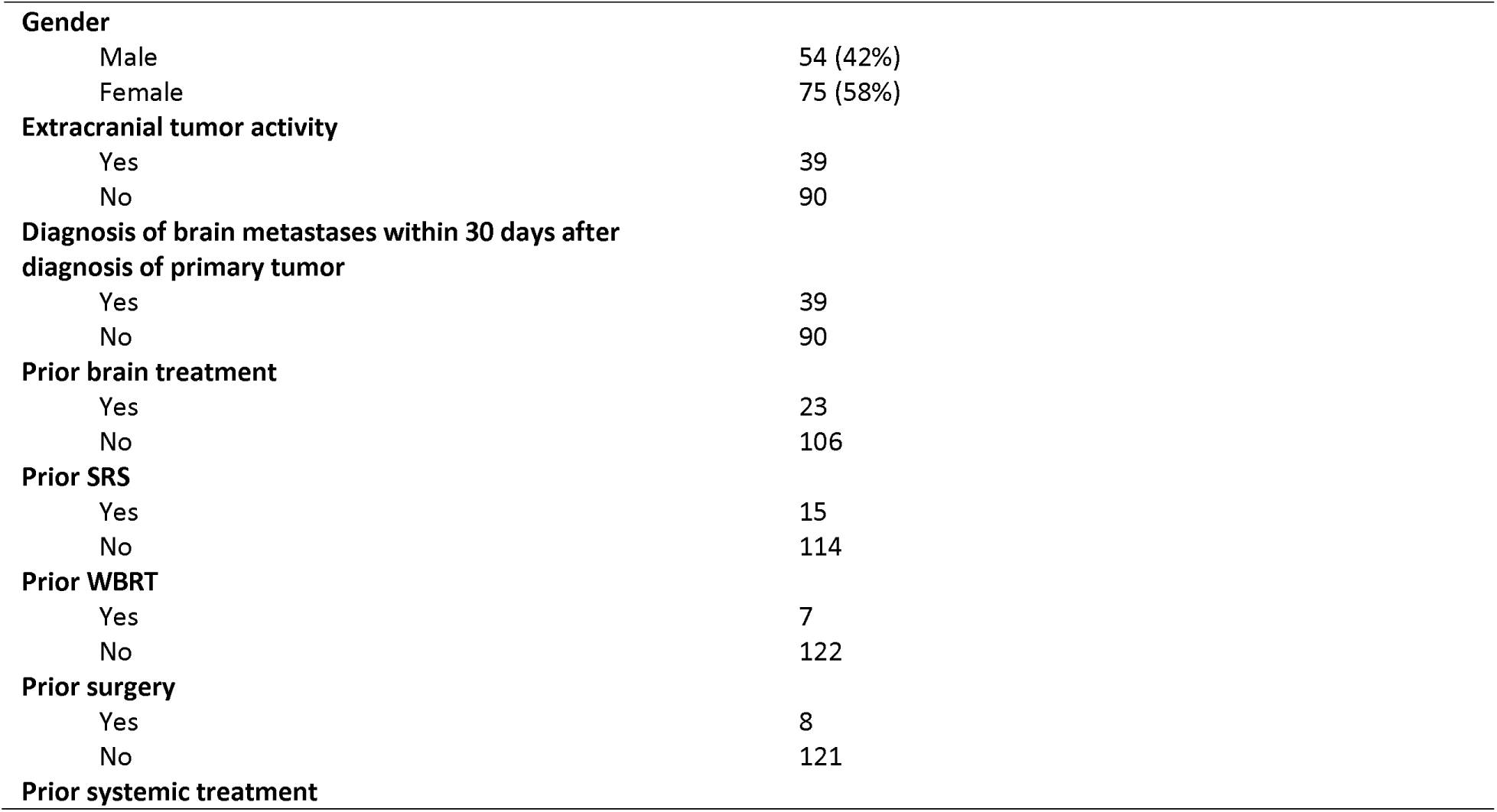
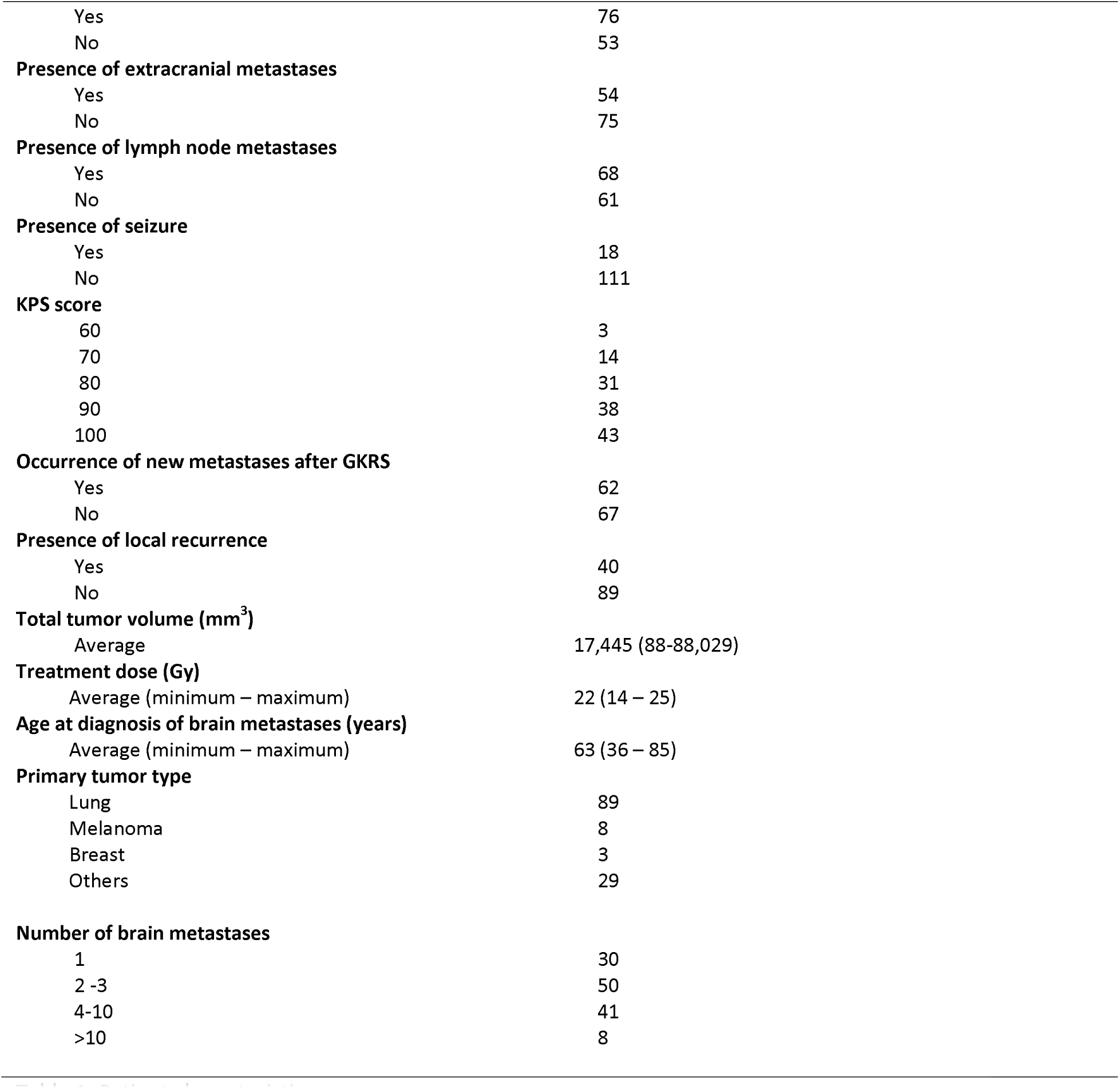
Patient characteristics.

The average performance metrics for the four models across the cross-validation datasets are shown in Table 2. In bold is the value of the best score for the corresponding performance metric.

**Table 2:**
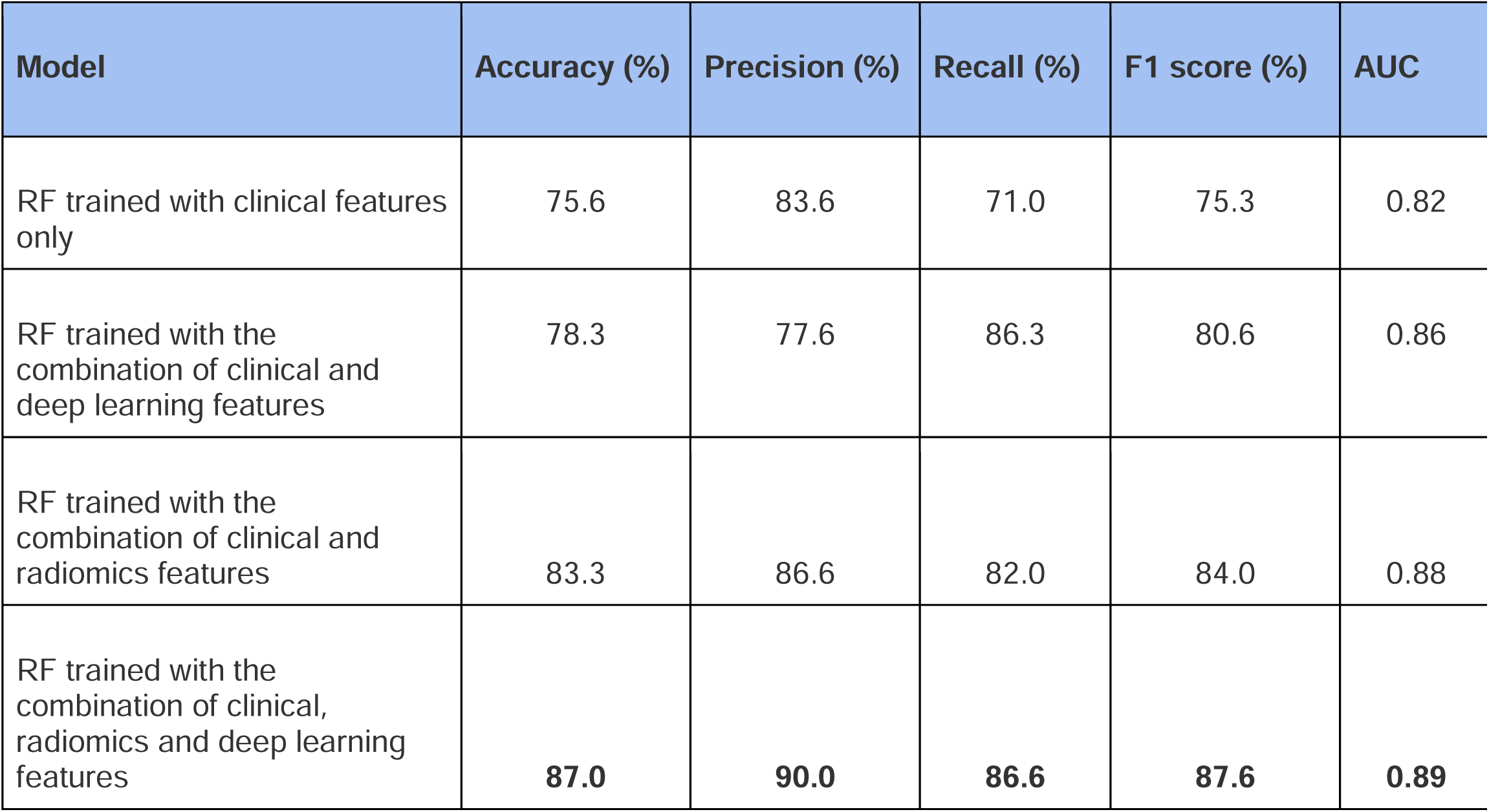
Average performance of the models on the validation datasets.

| Model | Accuracy (%) | Precision (%) | Recall (%) | F1 score (%) | AUC |
| --- | --- | --- | --- | --- | --- |
| RF trained with clinical features only | 75.6 | 83.6 | 71.0 | 75.3 | 0.82 |
| RF trained with the combination of clinical and deep learning features | 78.3 | 77.6 | 86.3 | 80.6 | 0.86 |
| RF trained with the combination of clinical and radiomics features | 83.3 | 86.6 | 82.0 | 84.0 | 0.88 |
| RF trained with the combination of clinical, radiomics and deep learning features | **87.0** | **90.0** | **86.6** | **87.6** | **0.89** |

The accuracy of the Random Forest (RF) model trained with clinical features only was 75.6%. The model trained with the combination of clinical and deep learning features had an improved accuracy of 78.3%. The model trained with the combination of clinical and radiomics features was even higher at 83.3%. But the model with the highest prediction accuracy of 87% was the model trained with the combination of clinical, radiomics and deep learning features. This model also achieved the highest precision of 90%, recall of 86.6%, F1 score of 87.6% and the best AUC of 0.89. The model trained with the combination of clinical and radiomics features showed the second highest accuracy of 83.3%, precision of 86.6%, F1 score of 84% and an AUC of 0.88. Interestingly, this model had a recall of 82% which is lower than the recall score (86.3%) of the model trained with the combination of clinical and deep learning features. However, the F1 score (which is the balance between the precision and recall) was higher for the model trained with clinical and radiomics features when compared with the model trained with the combination of clinical and deep learning features. The model with the lowest prediction accuracy of 75.6% was the model trained with clinical features only. This model also achieved the lowest recall of 71%, F1 score of 75.3% and the lowest AUC of 0.82. Though this model had better precision than the model trained with the combination of clinical and deep learning features, it still had a lower F1 score when compared to the model trained with the combination of clinical and deep learning features. The ROC curve of the model trained with the combination of clinical, radiomics and deep learning features is shown in Figure 3. The ROC curve of the other three models were added to the Appendix.

**Figure 3:**
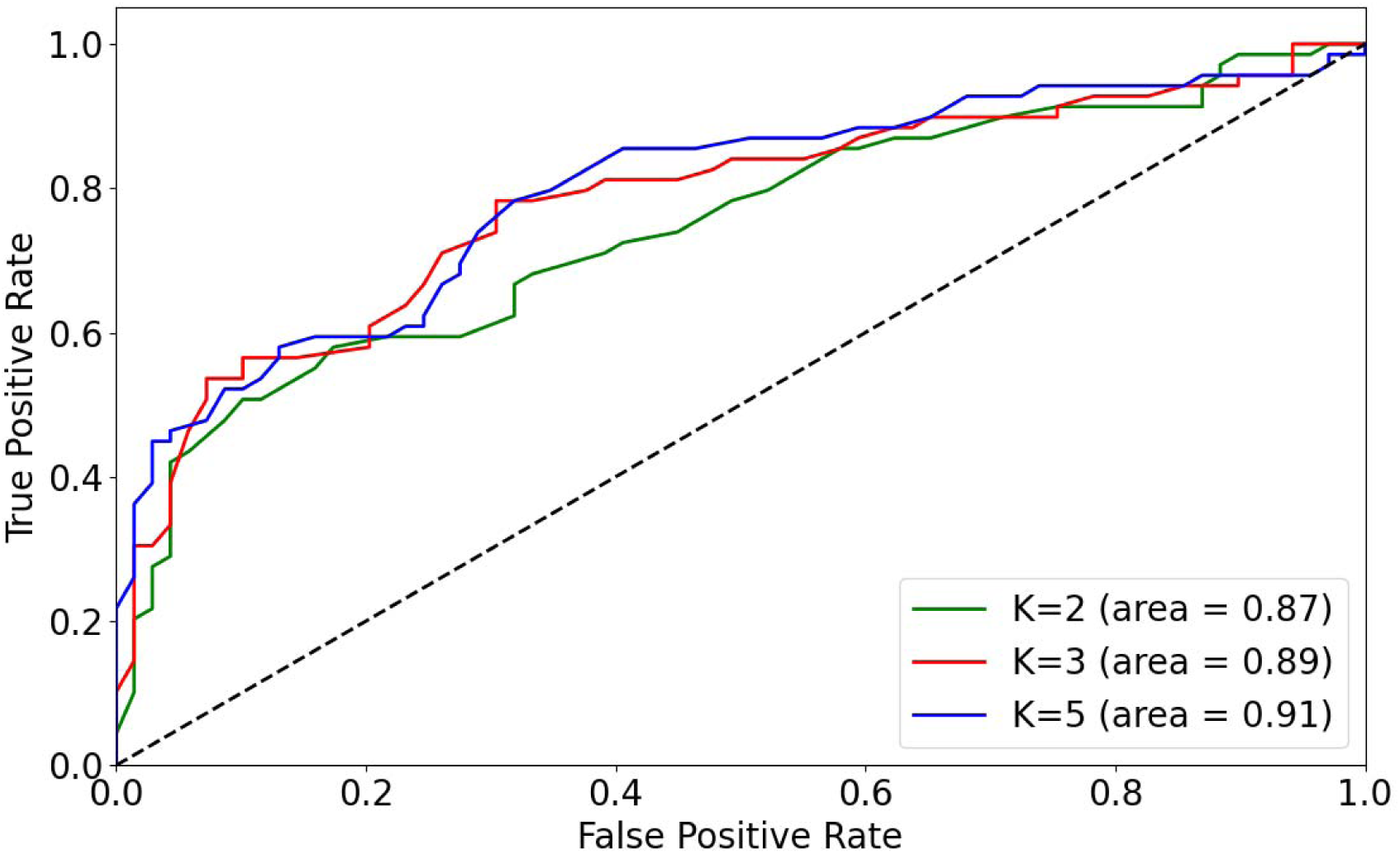
ROC curve of model trained with clinical, deep learning and radiomics features.

The variable importance of the top 10 factors for predicting the LC for each of the models were added to the appendix. For the model trained with the combination of clinical, radiomics and deep learning features, the top 10 features associated with the prediction of LC were original_glrlm_LowGrayLevelRunEmphasis, original_firstorder_Entropy, original_shape_LeastAxisLength, original_gldm_LargeDependenceHighGrayLevelEmphasis, original_shape_Elongation, original_firstorder_skewness, age at brain metastases diagnosis, original_shape_Maximum3DDiameter, age at primary tumor diagnosis, and original_glcm_SumSquares. Except for age at brain metastases diagnosis and age at primary tumor diagnosis which are clinical features, the rest of these top 10 features are radiomics features. There were no deep learning features in this list.

## Discussion

Brain metastases patients treated with SRT are at risk of developing local failure. Prompt diagnosis of local failure might increase treatment options and hence improve treatment outcomes. In this study, we trained and tested machine learning algorithms to predict the local failure using clinical features and T1-weighted MR imaging features. Four distinct models were developed: One model was trained with clinical features only, one with the combination of clinical and deep learning features, one with the combination of clinical and radiomics features and one with the combination of clinical, radiomics and deep learning features. Notably, the model incorporating all three feature sets outperformed the others, demonstrating the significance of integrating clinical, radiomics and deep learning features into predictive models.

Jalalifar et al [21] introduced a novel deep learning architecture to predict the outcome of LC in brain metastasis treated with stereotactic radiation therapy using treatment-planning magnetic resonance imaging and standard clinical attributes. The accuracy of the model developed with only the clinical features was 67.5%, but the addition of deep learning features to the clinical features increased the prediction accuracy to 82.5%. Our model trained with only the clinical and deep learning features provided a prediction accuracy of 78.3% (vs. 75.6% with clinical features only). Kawahara et al [28] proposed a neural network model using only the radiomics features for predicting the local response of metastatic brain tumor to GKRS. This proposed network provided a prediction accuracy of 78%. Karami et al [25] produced a radiomics framework to predict the outcome of SRT for brain metastases using the clinical and radiomics features. The framework predicted LC with an accuracy of 82%. Our model trained with only the clinical and radiomics features provided a prediction accuracy of 83.3%. To the best of our knowledge, the current study marks the first attempt to combine both radiomics and deep learning features with clinical features to predict LC of brain metastases. The model incorporating all these features predicted LC with an accuracy of 87%, surpassing all models trained with a subset of these features. This increased prediction accuracy holds promises for improved treatment outcomes for the patients. Furthermore, insights into the variable importances provided by the model could offer valuable insights into the features associated with the LC of brain metastases, potentially guiding future research and clinical decision-making.

The ability to predict LC with high accuracy before initiating SRT treatment offers an invaluable opportunity for tailoring treatment strategies for the best outcomes. Providing clinicians with information on the risk of local recurrence for individual patients empowers them to discuss these risks with patients prior to SRT. The capability to predict LC prior to treatment not only aids in informed decision-making regarding SRT but also opens avenues to consider alternative treatment modalities such as systemic therapy or WBRT. Additionally, it can enable clinicians to explore alternative radiotherapy approaches such as fractionated SRT or SRS with higher dose, depending on the predicted risk of local recurrence. Conversely, in cases where the risk of local recurrence is deemed low, SRT may be favored over other treatment options. Ultimately, pre-treatment prediction of LC serves as a valuable tool for both clinicians and patients, facilitating shared decision-making and optimizing treatment plans tailored to the needs and risk profiles of individual patients.

Although this study focused on creating a model for predicting the LC after SRT, the same approach can be extended to other treatment options and to the prediction of other clinical endpoints like overall survival.

One notable strength of this study lies in the meticulous brain metastases segmentation procedure. Expert oncologists and neuroradiologists at ETZ manually delineated the segmentations of the baseline GT on all the planning MRI scans used in this study. This ensures accurate regions of interest (ROIs) for the extraction of segment-based radiomics features.

It is important to note that in this study, we exclusively used the T1 weighted MRI scans. Exploring additional sequences and extracting radiomics and deep learning features from them could potentially improve the accuracy of the prediction models even more. In addition, for a more rigorous evaluation of the efficacy and robustness of the models, further investigations involving larger patient cohorts, preferably with multi-institutional data are warranted. Furthermore, the inclusion of an external validation dataset could significantly improve the generalizability of the prediction model, strengthening confidence in its clinical applicability across diverse patient populations and healthcare settings.

## Conclusion

The findings of this study show that the machine learning model trained with the combination of clinical, radiomics and deep learning features predict LC of brain metastases with high accuracy, outperforming models trained with the subset of these features. The increased prediction accuracy can lead to more tailored and effective interventions, resulting in improved treatment outcomes, prolonged patient survival, and enhanced quality of life.

## Data Availability

The data used for this study is available at ETZ and is accessible after approval from the ETZ Science office.

## Acknowledgments

We would like to acknowledge the support provided by Eline Verhaak for this research and thank her for helping us during the retrospective collection of clinical data from the Gamma Knife Center of the Elisabeth-TweeSteden Hospital at Tilburg, The Netherlands.

## Consent for publication

Not applicable.

## Contributions

All authors contributed to this research.

## Ethics approval

This study is part of the AI in Medical Imaging for novel Cancer User Support (AMICUS) project at Tilburg University. This project is approved by the Ethics Review Board at the Tilburg University.

## Consent to participate

The data did not contain any identifiable personal information, therefore the need for consent to participate was waived by the Institutional Review Board Elisabeth-TweeSteden Hospital (ETZ), Tilburg, The Netherlands (Study number : L1267.2021 - AMICUS).

## List of abbreviations

SRS: Stereotactic Radiosurgery
LC: Local Control
ETZ: Elisabeth-TweeSteden Hospital
AUC: Area Under the receiver operating characteristic Curve
WBRT: Whole-Brain Radiation Therapy
SRT: hypo-fractionated Stereotactic Radiotherapy
LF: Local Failure
AI: Artificial Intelligence
GKRS: Gamma Knife Radiosurgery
GT: Ground Truth
MRI: Magnetic Resonance Imaging
KPS: Karnofsky Performance Status score
ROC: Receiver Operating Characteristic
RF: Random Forest model
GPU: Graphical Processing Unit
ReLU: Rectified Linear Unit

## Appendix

### Full list of Radiomics features

original_shape_Elongation

original_shape_Flatness

original_shape_LeastAxisLength

original_shape_MajorAxisLength

original_shape_Maximum2DDiameterColumn

original_shape_Maximum2DDiameterRow

original_shape_Maximum2DDiameterSlice

original_shape_Maximum3DDiameter

original_shape_MeshVolume

original_shape_MinorAxisLength

original_shape_Sphericity

original_shape_SurfaceArea

original_shape_SurfaceVolumeRatio

original_shape_VoxelVolume

original_firstorder_10Percentile

original_firstorder_90Percentile

original_firstorder_Energy

original_firstorder_Entropy

original_firstorder_InterquartileRange

original_firstorder_Kurtosis

original_firstorder_Maximum

original_firstorder_MeanAbsoluteDeviation

original_firstorder_Mean

original_firstorder_Median

original_firstorder_Minimum

original_firstorder_Range

original_firstorder_RobustMeanAbsoluteDeviation

original_firstorder_RootMeanSquared

original_firstorder_Skewness

original_firstorder_TotalEnergy

original_firstorder_Uniformity

original_firstorder_Variance

original_glcm_Autocorrelation

original_glcm_ClusterProminence

original_glcm_ClusterShade

original_glcm_ClusterTendency

original_glcm_Contrast

original_glcm_Correlation

original_glcm_DifferenceAverage

original_glcm_DifferenceEntropy

original_glcm_DifferenceVariance

original_glcm_Id

original_glcm_Idm

original_glcm_Idmn

original_glcm_Idn

original_glcm_Imc1

original_glcm_Imc2

original_glcm_InverseVariance

original_glcm_JointAverage

original_glcm_JointEnergy

original_glcm_JointEntropy

original_glcm_MCC

original_glcm_MaximumProbability

original_glcm_SumAverage

original_glcm_SumEntropy

original_glcm_SumSquares

original_gldm_DependenceEntropy

original_gldm_DependenceNonUniformity

original_gldm_DependenceNonUniformityNormalized

original_gldm_DependenceVariance

original_gldm_GrayLevelNonUniformity

original_gldm_GrayLevelVariance

original_gldm_HighGrayLevelEmphasis

original_gldm_LargeDependenceEmphasis

original_gldm_LargeDependenceHighGrayLevelEmphasis

original_gldm_LargeDependenceLowGrayLevelEmphasis

original_gldm_LowGrayLevelEmphasis

original_gldm_SmallDependenceEmphasis

original_gldm_SmallDependenceHighGrayLevelEmphasis

original_gldm_SmallDependenceLowGrayLevelEmphasis

original_glrlm_GrayLevelNonUniformity

original_glrlm_GrayLevelNonUniformityNormalized

original_glrlm_GrayLevelVariance

original_glrlm_HighGrayLevelRunEmphasis

original_glrlm_LongRunEmphasis

original_glrlm_LongRunHighGrayLevelEmphasis

original_glrlm_LongRunLowGrayLevelEmphasis

original_glrlm_LowGrayLevelRunEmphasis

original_glrlm_RunEntropy

original_glrlm_RunLengthNonUniformity

original_glrlm_RunLengthNonUniformityNormalized

original_glrlm_RunPercentage

original_glrlm_RunVariance

original_glrlm_ShortRunEmphasis

original_glrlm_ShortRunHighGrayLevelEmphasis

original_glrlm_ShortRunLowGrayLevelEmphasis

original_glszm_GrayLevelNonUniformity

original_glszm_GrayLevelNonUniformityNormalized

original_glszm_GrayLevelVariance

original_glszm_HighGrayLevelZoneEmphasis

original_glszm_LargeAreaEmphasis

original_glszm_LargeAreaHighGrayLevelEmphasis

original_glszm_LargeAreaLowGrayLevelEmphasis

original_glszm_LowGrayLevelZoneEmphasis

original_glszm_SizeZoneNonUniformity

original_glszm_SizeZoneNonUniformityNormalized

original_glszm_SmallAreaEmphasis

original_glszm_SmallAreaHighGrayLevelEmphasis

original_glszm_SmallAreaLowGrayLevelEmphasis

original_glszm_ZoneEntropy

original_glszm_ZonePercentage

original_glszm_ZoneVariance

original_ngtdm_Busyness

original_ngtdm_Coarseness

original_ngtdm_Complexity

original_ngtdm_Contrast

original_ngtdm_Strength

### List of excluded features

original_firstorder_Uniformity

original_glcm_Id

original_glcm_Idm

original_glcm_Idmn

original_glcm_Idn

original_glcm_Imc2

original_glcm_InverseVariance

original_glcm_JointEnergy

original_glcm_MCC

original_glcm_MaximumProbability

original_gldm_LowGrayLevelEmphasis

original_gldm_SmallDependenceLowGrayLevelEmphasis

original_glrlm_GrayLevelNonUniformityNormalized

original_glrlm_LowGrayLevelRunEmphasis

original_glrlm_RunLengthNonUniformityNormalized

original_glrlm_RunPercentage

original_glrlm_ShortRunEmphasis

original_glrlm_ShortRunLowGrayLevelEmphasis

original_glszm_GrayLevelNonUniformityNormalized

original_glszm_LowGrayLevelZoneEmphasis

original_glszm_SmallAreaEmphasis

original_glszm_SmallAreaLowGrayLevelEmphasis

original_ngtdm_Coarseness

DL_5

DL_13

DL_31

DL_34

DL_43

DL_48

### Variable importance

**Figure 4.**
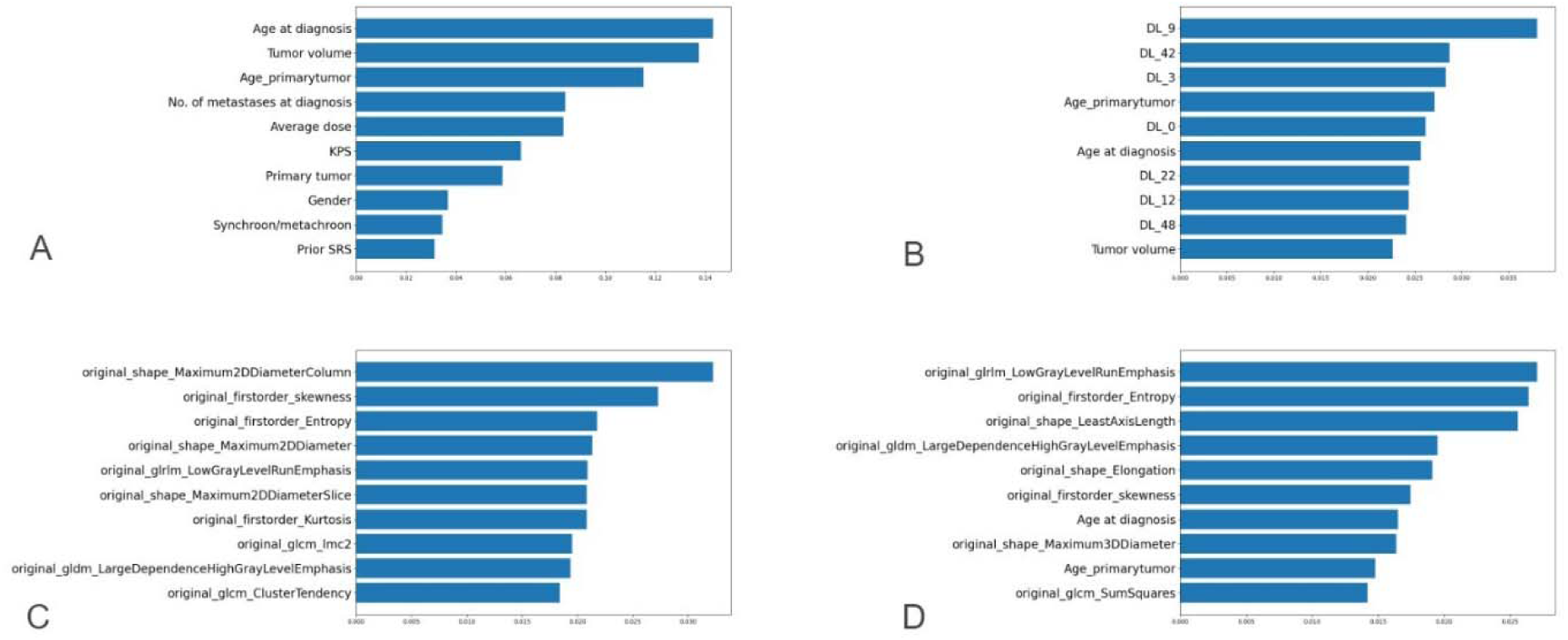
Variable importance for LC in decreasing order of significance for top 10 factors. A - model trained with clinical factors only. B - model trained with clinical and deep learning factors. C-model trained with clinical and radiomics features. D - model trained with the combination of clinical, radiomics and deep learning features.

### ROC Curves

**Figure 5:**
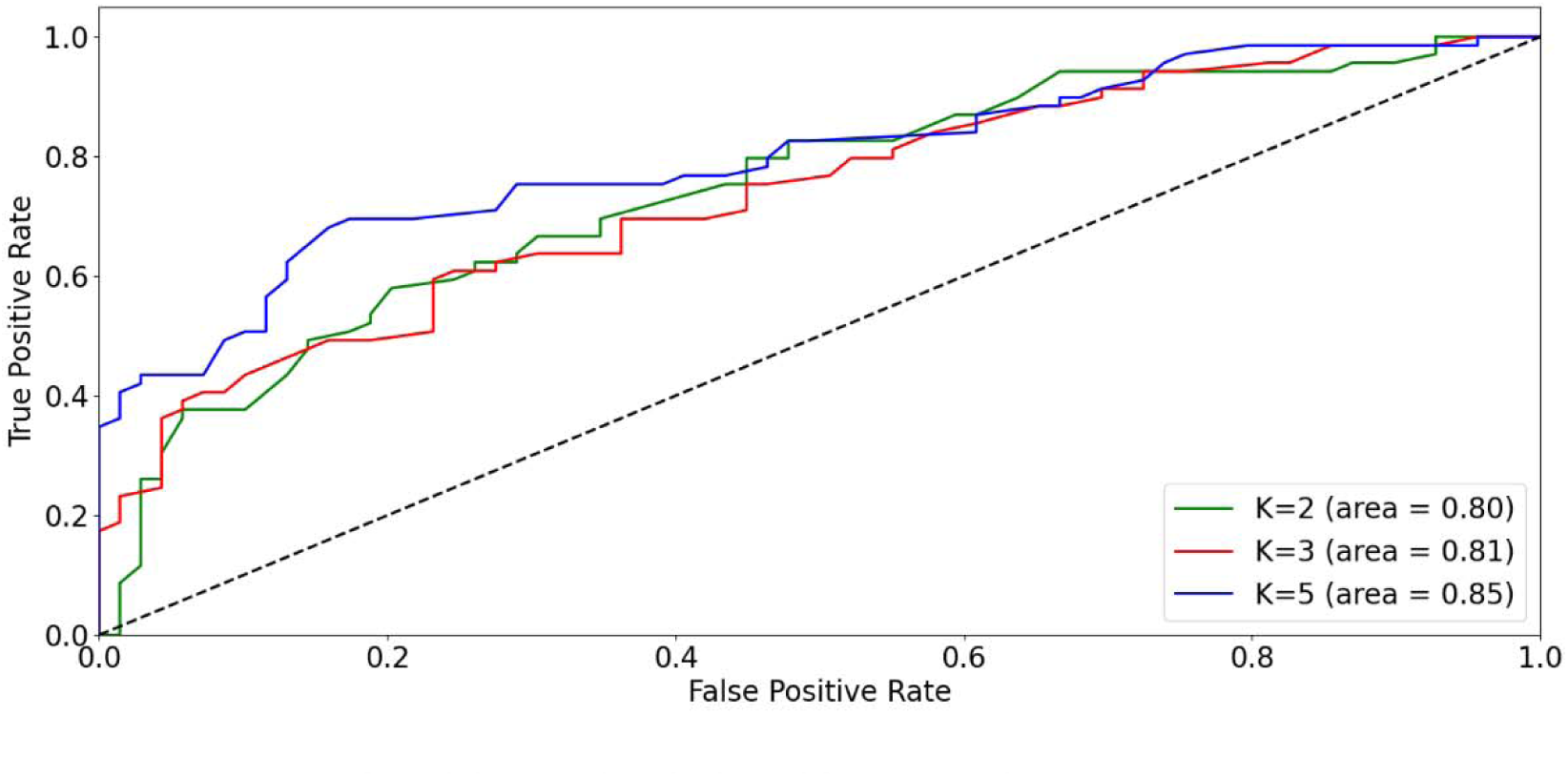
ROC curve of model trained with clinical features only.

**Figure 6:**
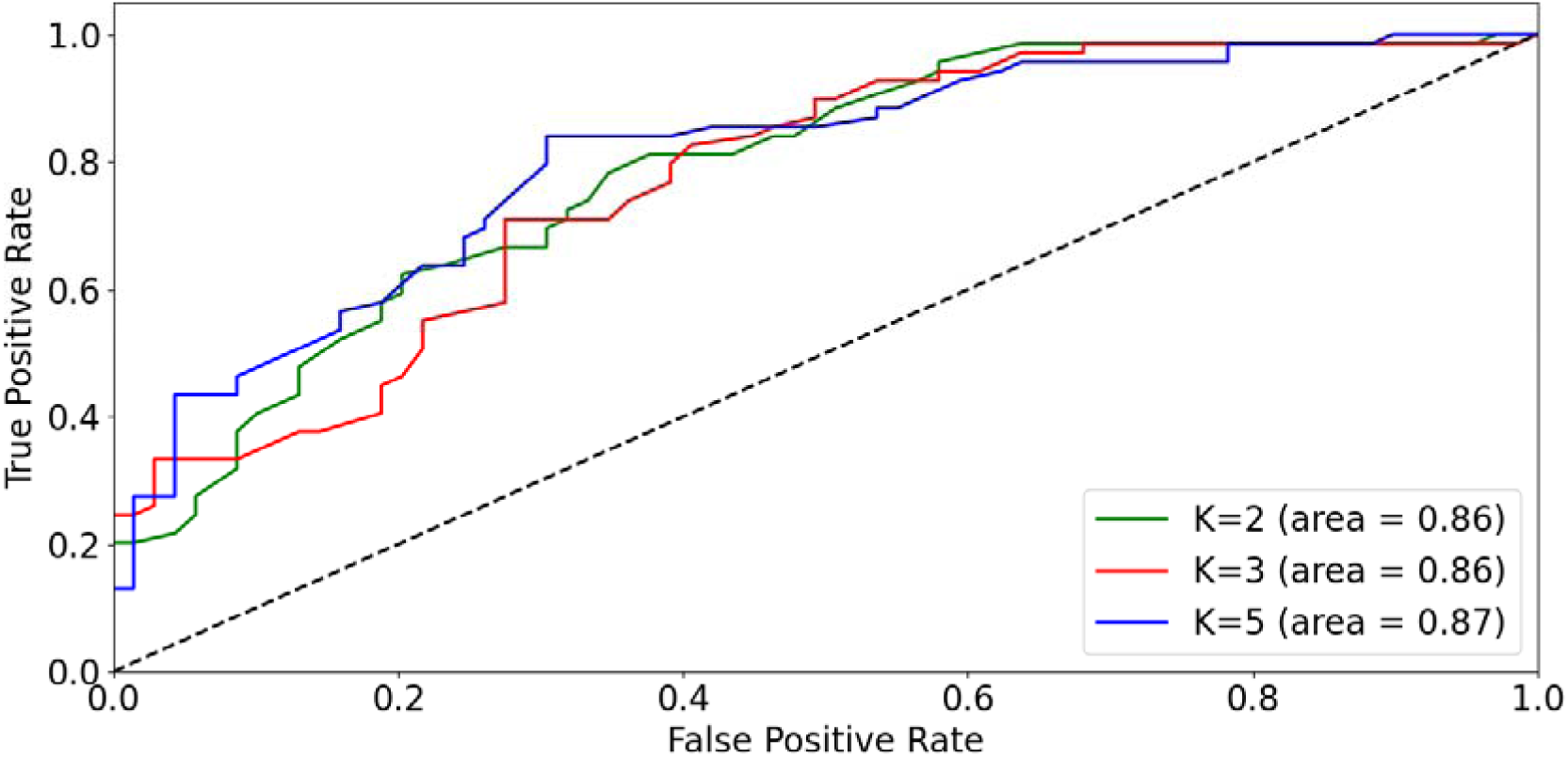
ROC curve of model trained with clinical and deep learning features.

**Figure 7:**
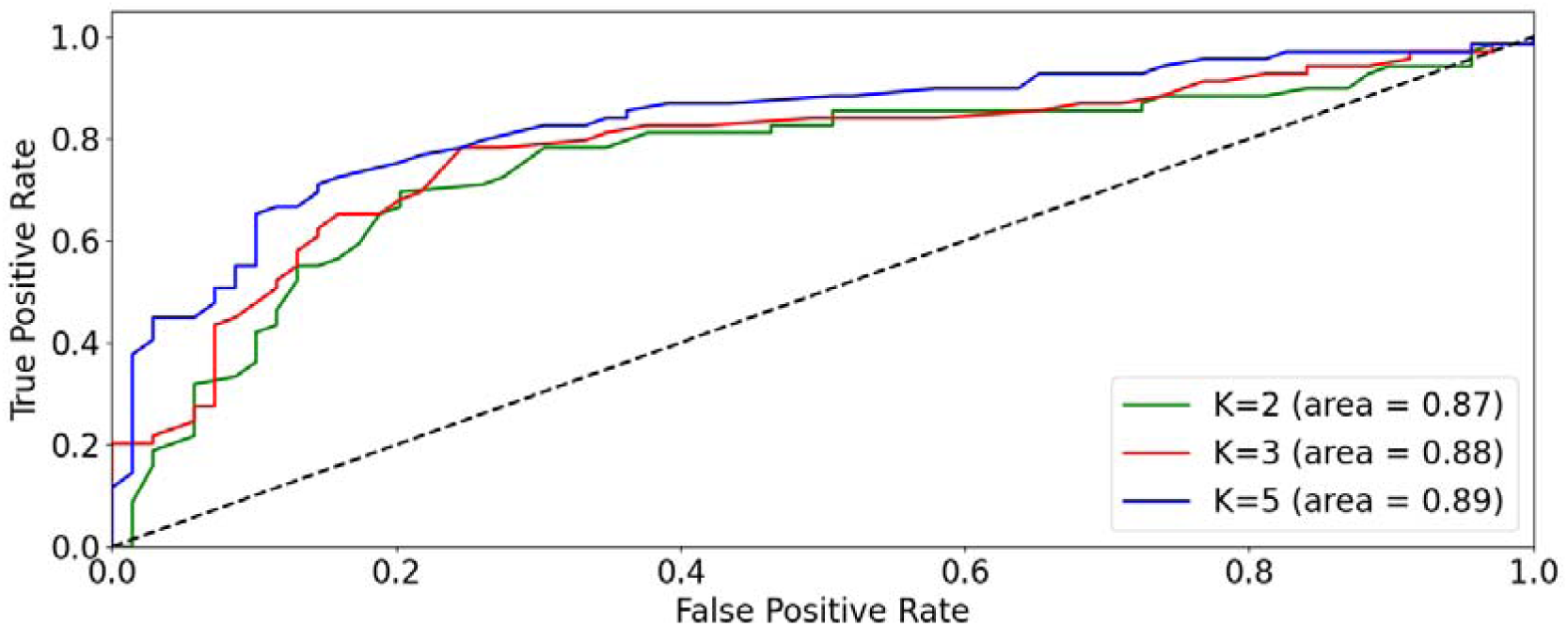
ROC curve of model trained with clinical and radiomics features.

